# Do mass-spectrometry-derived metabolomics improve prediction of pregnancy-related disorders? Findings from a UK birth cohort with independent validation

**DOI:** 10.1101/2021.05.04.21256218

**Authors:** Nancy McBride, Paul Yousefi, Ulla Sovio, Kurt Taylor, Yassaman Vafai, Tiffany Yang, Bo Hou, Matthew Suderman, Caroline Relton, Gordon C. Smith, Deborah A. Lawlor

## Abstract

Maternal risk factors, such as body mass index (BMI), age, smoking, parity and ethnicity, are associated with risk of pregnancy-related disorders. However, many women who experience gestational diabetes (GDM), gestational hypertension (GHT), pre-eclampsia (PE), have a spontaneous preterm birth (sPTB) or an offspring born small/large for gestational age (SGA/LGA), do not display any of these risk factors. Tools that better predict these outcomes are needed to tailor antenatal care to risk. Recent studies have suggested that metabolomics may improve the prediction of these pregnancy-related disorders. These have largely been based on targeted platforms or focused on a single pregnancy outcome. The aim of this study was to assess the predictive ability of an untargeted platform of over 700 metabolites to predict the above pregnancy-related disorders in two cohorts.

We used data collected from women in the Born in Bradford study (BiB; two sub-samples, n=2,000 and n=1,000) and the Pregnancy Outcome Prediction study (POPs; n=827) to train, test and validate prediction models for GDM, PE, GHT, SGA, LGA and sPTB. We used ten-fold cross-validation and penalised regression to create prediction models. We compared the predictive performance of 3 models: 1) risk factors (maternal age, pregnancy smoking, BMI, ethnicity, and parity) 2) mass spectrometry (MS)-derived metabolites (N = 718 quantified metabolites, collected at 26-28 weeks’ gestation) and 3) combined risk factors and metabolites. We used BiB for training and testing the models and POPs for independent validation.

In both cohorts, discrimination for GDM, PE, LGA and SGA improved with the addition of metabolites to the risk factor model (combined risk factor and metabolite model). The combined models’ area under the curve (AUC) were similar for both cohorts, with good discrimination for GDM (AUC (95% CI) BiB 0.76 (0.71,0.81) and POPs 0.76 (0.72,0.81)) and LGA (BiB 0.86 (0.80,0.91) and POPs 0.76 (0.60,0.92)). Discrimination was improved for the combined models (compared to the risk factors models) for PE and SGA, with modest discrimination in both studies (PE - BiB 0.68 (0.58,0.78) and POPs 0.66 (0.60,0.71); SGA - BiB 0.68 (0.63,0.74) and POPs 0.64 (0.59,0.69)). Prediction for sPTB was poor in BiB and POPs for all models, with AUC ∼0.5. In BiB, calibration for the combined models was good for GDM, LGA and SGA. Retained predictors include 4-hydroxyglutamate for GDM, LGA and PE, and glycerol for GDM and PE.

MS-derived metabolomics combined with maternal risk factors improve prediction of GDM, PE, LGA and SGA, with good discrimination for GDM and LGA. Validation across two very different cohorts supports further investigation on whether the metabolites reflect novel causal paths to GDM and LGA. Developing these prediction tools could enable tailoring antenatal care to improve earlier and more accurate identification of high-risk women.

## Introduction

Gestational diabetes (GDM), gestational hypertension (GHT), pre-eclampsia (PE), small for gestational age (SGA), large for gestational age (LGA) and spontaneous preterm birth (sPTB) are common pregnancy-related disorders(1-7) - associated with long term mortality and morbidity in mother and offspring(7-10). Currently, prediction of these disorders relies largely on stratifying women based on established risk factors. Established risk factors for predicting these disorders include maternal smoking(11), age(12), ethnicity(13), parity(14) and body mass index (BMI)(15). However, many women do not meet any of these risk factors and yet go on to have a complicated pregnancy(16-18). A good indicator of risk is previous pregnancy history, however this is not obtainable in nulliparous women(19, 20) and there is a need for clinical prediction models that do not depend on previous pregnancy history(19). Development of such might result in better ways of managing antenatal care by intensely monitoring higher risk women and avoiding unnecessary intervention in low-risk women.

Pregnancy is characterised by widespread metabolic changes(21-23). Metabolomics, the quantification of molecules arising from metabolic processes, could improve the prediction of common pregnancy-related disorders(24). We have recently shown that a targeted nuclear magnetic resonance (NMR)-derived metabolomics panel of 156 (mostly lipid) traits, can improve prediction of GDM, LGA and hypertensive disorders of pregnancy (HDP) in Born in Bradford (BiB), a large general population pregnancy cohort. This work was externally validated in the UK Pregnancies Better Eating and Activity Trial (UPBEAT), a cohort of obese pregnant women(25). We have also identified novel metabolite predictors for specific pregnancy outcomes using a mass spectrometry (MS)-derived metabolites platform in the Pregnancy Outcome Prediction study (POPs) which were externally validated in the BiB cohorts. Specifically, we found that the amino acid 4-hydroxyglutamate improves prediction of PE in term pregnancy. A ratio of the product of the plasmalogen 1-(1-enyl-stearoyl)-2-oleoyl-GPC (P-18:0/18:1) and the steroid 5α-androstan-3α,17α-diol disulfate to the product of the carbohydrate 1,5-anhydroglucitol and the polyamine N1,N12-diacetylspermine was a better predictor of fetal growth restriction/SGA than the clinically validated biomarker soluble fms-like tyrosine kinase 1 and placental growth factor ratio (sFlt:P lGF)(26, 27). In the previous studies we used serial samples from POPs and the focus was identifying a small number of independently predictive metabolites which might form the basis of a targeted assay. Here, we use the entire metabolomic profile in a high dimensional analysis, developed in one cohort and validated in another.

To date, most studies have focused on a narrow range of metabolites and/or on a single pregnancy outcome, lack external validation and suffer from over fitting. The aim of this study is to determine whether metabolites included in an extensive MS metabolomics platform can improve prediction of six common pregnancy outcomes (GDM, PE, GHT, SGA, LGA and sPTB) over established risk factors alone. Here, we validate the results externally and show metabolomics enhances prediction over maternal characteristics risk factors alone. The multimorbidity that exists between these disorders may mean a prediction tool for more than one could be clinically valuable. We therefore compare overlap between traits across the prediction models for each outcome, and whether a single prediction model can be developed to predict the occurrence of any of the six outcomes.

## Method

### Participants

BiB is a population-based prospective birth cohort that recruited 12,453 women who had 13,776 pregnancies. Full details of the study methodology were reported previously(28). In brief, most women were recruited at their oral glucose tolerance test (OGTT) at approximately 26–28 weeks gestation, which is offered to all women booked for delivery at Bradford Royal Infirmary, except those with known diabetes. Eligible women had an expected delivery between March 2007 and December 2010. Bradford, in the North of England, is one of the most deprived cities in the UK. In BiB, most of the obstetric population consists of women of white British or Pakistani origin (together accounting for 81%). Ethical approval for the study was granted by the Bradford National Health Service Research Ethics Committee (ref 06/Q1202/48).

The BiB metabolomics data has been described in detail previously in a data note(29). In brief, 3,000 women were selected for plasma MS metabolomic profiling using samples taken at 26-28 weeks gestation(28). These 3,000 women had profiling in two separate sub-samples of 1,000 women and 2,000 women. There was no participant overlap. Dataset 1 was completed in December 2017 and consisted of 1,000 mother (pregnancy) and offspring (cord blood) pairs. These were randomly sampled from pairs where both had a suitable sample for analyses and belonged to one of the two main ethnic groups in BiB (Pakistani or White British) (**Figure 1** and **Figure S1a**). Only the maternal pregnancy metabolites are used in this study. Dataset 1 will be referred to as “BiB 1,000”. Dataset 2 (“BiB 2,000”) was completed in December 2018 and consisted of 2,000 mothers (**Figure 1** and **Figure S1b**) selected using a case-cohort design, including cases of GDM, PE, GHT, SGA, LGA, PTB, stillbirths and congenital anomalies, together with a randomly selected sub-cohort of the whole eligible cohort (30). BiB 2,000 was used for training the prediction models, and the BiB 1,000 for testing them.

**Figure 1.**
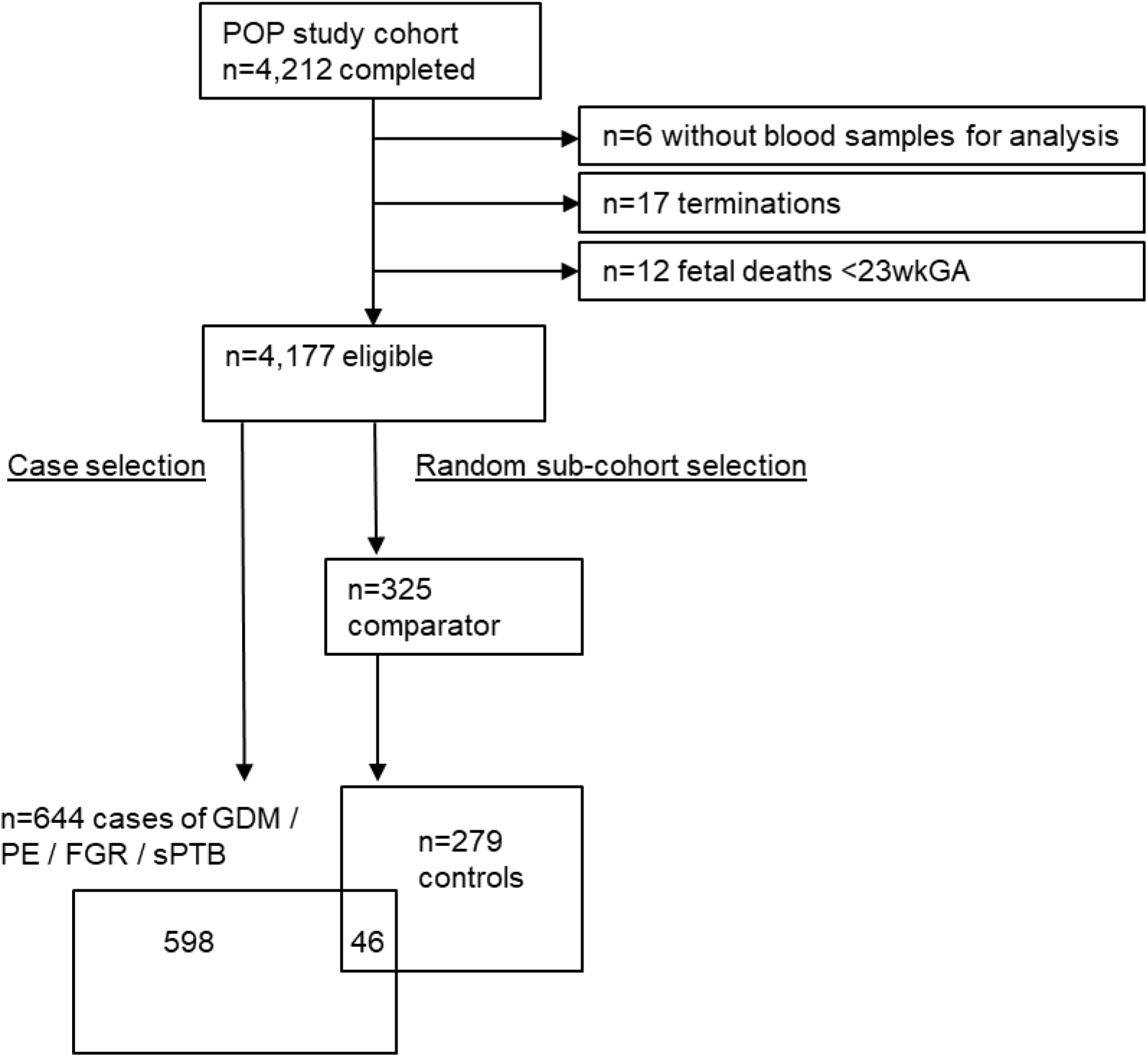
Illustrating the flow of participants into the Metabolon datasets (**1a:** BiB 1,000 and **1b:** BiB 2,000 and **1c** POPs (N = 923) cohorts). Abbreviations: MS, mass spectrometry; BiB, Born in Bradford; GWAS, genome wide association study; EDTA, ethylenediamine tetraacetic acid; HDP, hypertensive disorders of pregnancy; GDM, gestational diabetes; GHT, gestational hypertension; PE, pre-eclampsia, PTB, preterm birth; sPTB, spontaneous preterm birth; CA, congenital anomaly; SB, still birth; FGR, fetal growth restriction; GA, gestational age. Figures 1A and 1B taken from our data note(29), with permission.

**Figure 1A and 1B:**
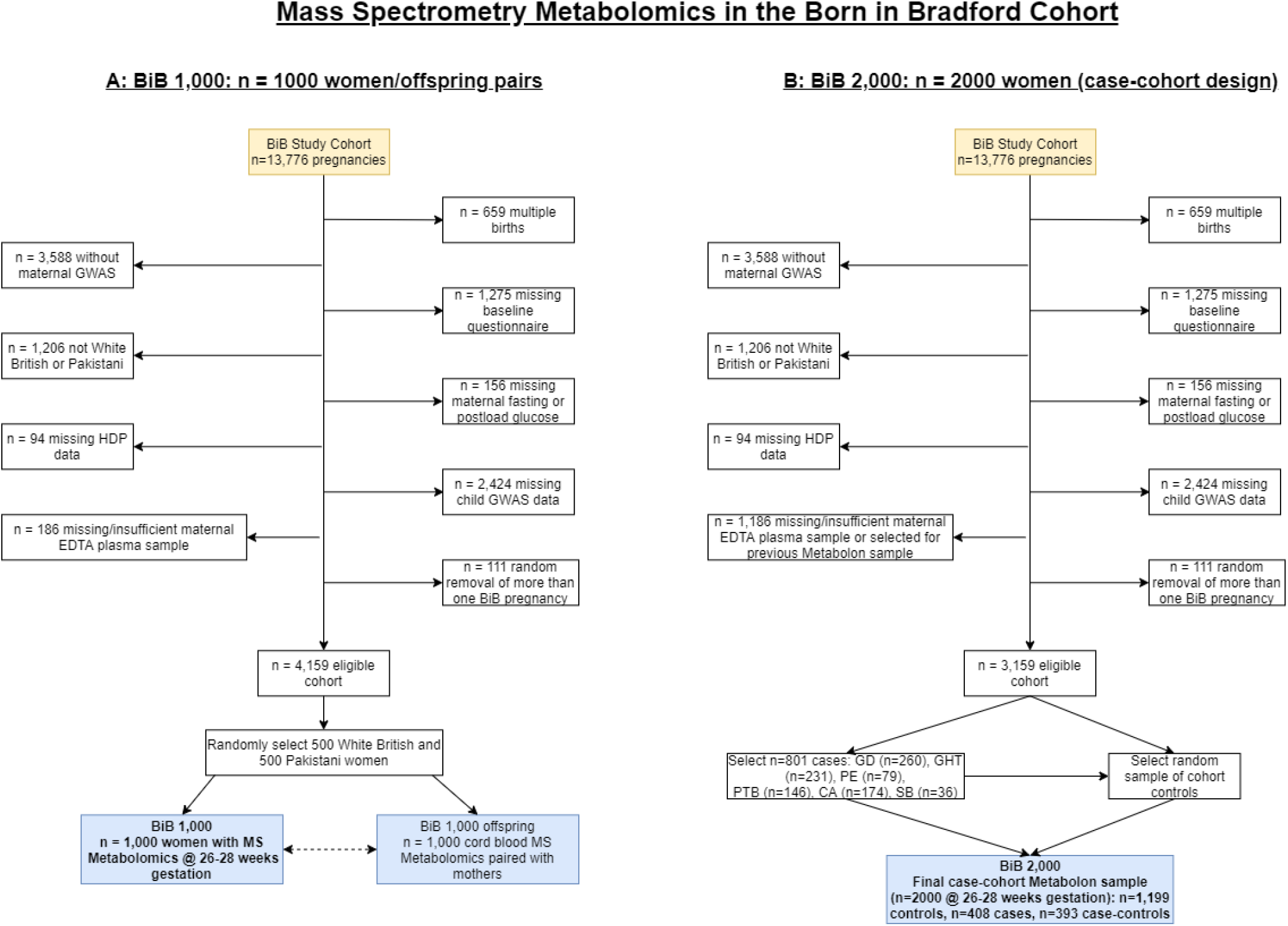
Born in Bradford flowchart: the selection of participants for mass spectrometry metabolomic profiling in the Born in Bradford 1,000 (A) and 2,000 (B). Taken with permissions from Taylor & McBride et al. Abbreviations: MS, mass spectrometry; BiB, Born in Bradford; GWAS, genome wide association study; EDTA, ethylenediaminetetraacetic acid; HDP, hypertensive disorders of pregnancy; GD, gestational diabetes; GHT, gestational hypertension; PE, pre-eclampsia, PTB, preterm birth; CA, congenital anomaly; SB, still birth.

External validation was undertaken in POPs, a prospective cohort study of unselected nulliparous women(19). Those eligible were recruited at the Rosie Hospital, Cambridge, U.K., between January 2008 and July 2012. Cambridge is an affluent city in Eastern England, and participants in POPs are nulliparous and mostly of White European origin. Ethical approval was obtained from the Cambridgeshire Research Ethics Committee (reference number 07/H0308/163). All study participants gave written informed consent. POPs also utilise a case-cohort design, previously described elsewhere (**Figure S2**)(19, 30). Only singleton pregnancies were included in this study.

### Metabolomic predictors

The untargeted MS analysis of BiB and POPs samples was performed at Metabolon, Inc. (Durham, North Carolina, USA) using a modification of a previously described ultra-performance liquid chromatography (UPLC) mass spectrometry (MS) method (UPLC-MS/MS)(27, 30). The platform consisted of four independent UPLC-MS/MS methods. This method provides relative quantification of >1,000 metabolites in key classes: amino acids, carbohydrates, lipids, nucleotides, carbon metabolism, vitamins, xenobiotics, and novel unidentified metabolites. Metabolite concentrations were quantified using area-under-the-curve of primary MS ions and were expressed as the means of the medians (MoM) value for all batches processed on the given day. Metabolon data are provided in a quantified (scaled) data set. Samples from the BiB 1,000, BiB 2,000 and POPs were run in different batches and are all analysed separately in this study (BiB 2,000 for training, BiB 1,000 for testing, and POPs for external validation). In BiB, MS analyses were undertaken on ethylenediamine tetraacetic acid (EDTA) plasma taken around 26-28 weeks gestation. In POPs, MS analyses were undertaken on maternal serum samples from 12, 20, 28 and 36 weeks of gestation. In this study, we used the 28-week gestation timepoint for our external validation as this best matched the gestation of the BiB samples. Previous work has shown that reproducibility in both serum and plasma is good. If the same sample procedures are used, either matrix should yield similar results(31). Further information on the metabolomic data in both cohorts have been published previously(29, 30, 32). The participant selection workflows are available in **Figures S1a, S1b** and **S1c**. We included 718 metabolites in our models, which is the number of metabolites available in BiB 1,000, BiB 2,000 and POPs. A full list of the metabolites and their pathways included in this study as predictors is provided in **Table S1**.

### Risk factor predictors

We compared metabolomic prediction models to models of risk factor predictors that are routinely collected in antenatal care: maternal age, early-pregnancy BMI, parity, ethnicity, and smoking during pregnancy. This information was collected during recruitment or extracted from clinical records. In BiB, data on parity and weight were extracted from the first antenatal clinic records (around 12-weeks of gestation). Weight (kg, Seca 2in1 scales, Harlow Healthcare Ltd, London, UK) and height (cm) were measured using established protocols at recruitment and used to calculate BMI. Parity was dichotomized as having experienced one or more previous pregnancy ≥24 weeks gestation, or no previous pregnancy. Data on age, ethnicity, and smoking were obtained from questionnaires administered by the BiB research fieldworkers at recruitment. These fieldworkers included people fluent in the common languages used by women undergoing antenatal care in Bradford at the time of recruitment, including Urdu and Mirpuri. In BiB, ethnicity was self-reported or obtained from primary care medical records and categorised as either White British or Pakistani (the only two ethnic groups who had metabolomic profiling, and who together include over 85% of the BiB mothers).

In POPs, only nulliparous women were recruited(33-35). Weight measured at recruitment (∼12 weeks) was used for the calculation of BMI and maternal age was defined as age at recruitment. Maternal height was measured at ∼20 weeks. Maternal ethnicity and smoking were self-reported using a questionnaire at ∼20 weeks in POPs, and ethnicity was categorised as White or non-White for the present analysis. In both studies, smoking was dichotomised as any smoking during pregnancy.

### Outcomes

We examined predictive discrimination for six pregnancy-related disorders: GDM, GHT, PE, SGA, LGA and sPTB. In BiB, blood pressure and proteinuria measurements taken at any time during pregnancy were extracted from medical records. In BiB, GHT was defined as new onset of elevated blood pressure (systolic blood pressure ≥140 mmHg or greater, and/or diastolic blood pressure ≥90 mmHg or greater) after 20 weeks’ gestation on two or more occasions. In POPs, cases of GHT were not identified for inclusion in the case-cohort sub-sample on whom metabolites were assayed. Consequently, we were unable to explore external validation of our GHT prediction models. In BiB, PE was defined as GHT plus clinically significant proteinuria, defined as 1 or greater ‘+’ on the reagent strip reading (equivalent to 30mg/mmol) or greater on spot urine protein/creatinine ratio (4). In POPs, PE was defined according to the 2013 American College of Obstetricians and Gynaecologists (ACOG) guidelines(30, 36). The selection of cases did not include non-severe superimposed PE associated with delivery at term, meaning PE in POPs likely reflects a somewhat more severe group than in BiB. In BiB, GDM was defined according to modified World Health Organization (WHO) definition used in clinical practice at the time; fasting glucose ≥ 6.1 mmol/L or 2hr post-load glucose ≥ 7.8 mmol/l(5). In POPs, between 2008 and 2010, GDM diagnosis was based on diagnostic criteria adapted from the WHO recommendations: fasting, 1hr, and 2hr glucose levels ≥ 6.1 mmol/L, ≥ 10.0 mmol/L or ≥ 7.8 mmol/L, respectively. From 2011 onwards, these were replaced locally with diagnostic criteria adapted from the International Association of Diabetes and Pregnancy Study Groups’ recommendations: fasting, 1hr, and 2hr glucose levels ≥ 5.3 mmol/L, ≥ 10.0 mmol/L, or ≥ 8.5 mmol/L, respectively(37).

In both studies, we used the Hadlock formula to derive gestational age and sex standardised birthweight percentiles as previously described (38, 39). SGA was defined as <5 ^th^ percentile birth weight (being a more accurate measure of fetal growth restriction than the conventional 10^th^ percentile) and LGA as ≥90^th^ percentile birthweight. In both cohorts, sPTB was defined as spontaneous onset of labour (i.e., without medical or surgical induction or elective Caesarean section) before 37 completed weeks of gestation. In POPs, there was additional criteria that delivery occurred after 24 completed weeks of gestation. Whilst this specific criterion was not applied to BiB, no births before 24 weeks gestation were included in the metabolomic profiling.

### Statistical analysis

All statistical analysis was performed in R 3.5.1, R.1.3.1 or STATA 15.1. In POPs, prior to this paper’s analysis, one woman’s BMI was imputed by sample mean and 15 women’s ethnicity was imputed by the most common category (White), since the proportion of missing values in these variables was very small. Aside from these imputations, all data were complete, and any participants with missing data on risk factors or outcomes were excluded from the analysis.

### Comparison groups in the case-cohort BiB 2000 and POPs

BiB 2,000 and POPs sampled participants for metabolomic analyses using a case-cohort design which increases statistical power (by including all cases for each outcome). This allows the analysis of associations of metabolites with multiple outcomes with the comparison group for each cohort being the equivalent of a random sample of the underlying cohort(40). In a case-cohort design, we only use the non-cases of the given outcome from the random sub-cohort as controls. Therefore, we remove oversampled ‘cases’ that are not a case of interest for this study.

### Main analyses

Only metabolites present in all three datasets (BiB 1,000, 2,000 and POPs) were included in the prediction models (n=718). Prior to analysis, scaled and imputed metabolite values were log transformed and then converted to standard deviation (SD) units by subtracting the sample mean from their logged value and dividing by the sample SD of logged values, within the prediction model. This results in some metabolites with very little variance. We applied a variance threshold in BiB of more than 440 unique values. This reduced the number of metabolites in the dataset from 1,371 to 1,074 in the BiB 1,000 and 1,241 to 1,152 in the BiB 2,000. We then only included metabolites present in all three datasets (BiB 1,000, 2,000 and POPs) and included these in the prediction models (n=718). In the training dataset (BiB 2,000), we optimized elastic net penalized regression fit by tuning model hyperparameters, alpha and lambda, using 10-fold cross validation implemented with the *caret* package in R(41). Optimal parameter values were selected that minimised cross validated error in the training set. The lambda parameter constrains the sum of the individual predictor coefficient values, performing feature selection by ‘shrinking’ coefficients on non-informative predictors to zero and therefore removing them from the model. Out-of-sample prediction performance was further assessed by AUC and calibration slopes in both the BIB 1,000 test set and external validation in POPs. In all, we assessed ability to discriminate pregnancy-related disorder by comparing AUC values for three models: 1) established risk factors model, 2) MS-derived metabolites model and 3) combined established risk factors and metabolites model.

### Additional analysis

When studying preterm birth, POPs focused on those cases which were due to spontaneous preterm labour (sPTB). For concordance, sPTB was also used in BiB, despite the availability of cases of non-spontaneous preterm birth. However, in an additional analysis, we also trained and tested a model for predicting any PTB, defined as any delivery < 37 completed weeks in the BiB cohorts, irrespective of the onset of labour.

We wanted to see whether we could create an accurate prediction model for ‘any’ pregnancy-related disorder. We created a binary variable of ‘any’ of GDM, GHT, PE, LGA, SGA or sPTB cases, compared with women who had experienced none of those. As in the main analysis, for these additional analyses we trained the models in BiB 2,000 and tested them in BiB 1,000, assessed discrimination using AUC and calibration using calibration slopes.

## Results

The BiB 1,000 and BiB 2,000 have similar distributions of age, BMI, parity, smoking and, by design, both are around half White British and half Pakistani (**Table 1**). In contrast, around 95% of the POPs women studied are of White ethnicity. The BiB 1,000 and BiB 2,000 have higher smoking prevalence than POPs. BiB is a cohort of both multi and nulliparous women, whereas POPs is all nulliparous women.

**Table 1:**
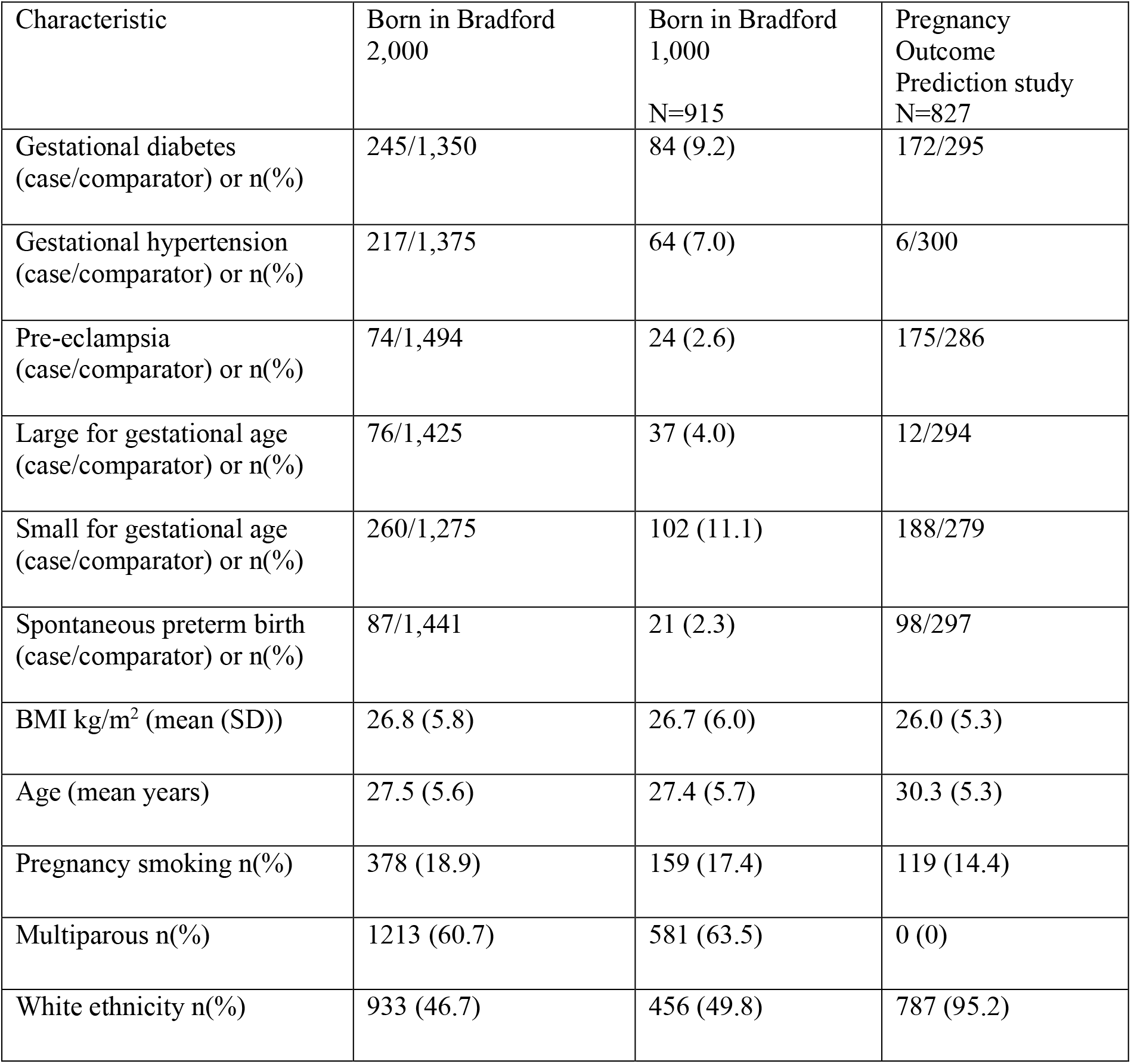
Participant characteristics from the three participating cohorts: Born in Bradford 2,000, Born in Bradford 1,000 and the Pregnancy Outcome Prediction study. Data in this table are complete. BiB 2,000 and POPs used a case cohort design, i.e. they were over-sampled for cases. In these two studies, the total numbers vary depending on the outcome. For the distributions of risk factor predictors in this table, we have used the overall mass spectrometry sample cohorts, n=2,000 for BiB 2,000 and n=827 for POPs (Figure S1). Because of substantial oversampling of cases in these studies, we do not give a prevalence (%) for the outcomes but rather give the numbers of cases and number in the comparison group for each outcome. The number of women in the comparator group varies per outcome as some from the comparator group are always relabelled as cases. POPs did not have an adequate number of women with GHT, hence no validation analysis was performed. Abbreviations: BMI, body mass index.

| Characteristic | Born in Bradford<br>2,000 | Born in Bradford<br>1,000<br>N=915 | Pregnancy<br>Outcome<br>Prediction study<br>N=827 |
| --- | --- | --- | --- |
| Gestational diabetes<br>(case/comparator) or n(%) | 245/1,350 | 84 (9.2) | 172/295 |
| Gestational hypertension<br>(case/comparator) or n(%) | 217/1,375 | 64 (7.0) | 6/300 |
| Pre-eclampsia<br>(case/comparator) or n(%) | 74/1,494 | 24 (2.6) | 175/286 |
| Large for gestational age<br>(case/comparator) or n(%) | 76/1,425 | 37 (4.0) | 12/294 |
| Small for gestational age<br>(case/comparator) or n(%) | 260/1,275 | 102 (11.1) | 188/279 |
| Spontaneous preterm birth<br>(case/comparator) or n(%) | 87/1,441 | 21 (2.3) | 98/297 |
| BMI kg/m<sup>2</sup> (mean (SD)) | 26.8 (5.8) | 26.7 (6.0) | 26.0 (5.3) |
| Age (mean years) | 27.5 (5.6) | 27.4 (5.7) | 30.3 (5.3) |
| Pregnancy smoking n(%) | 378 (18.9) | 159 (17.4) | 119 (14.4) |
| Multiparous n(%) | 1213 (60.7) | 581 (63.5) | 0 (0) |
| White ethnicity n(%) | 933 (46.7) | 456 (49.8) | 787 (95.2) |

### Predictors retained in the models

**Table 2** shows the number of predictors retained in each model during model training in BiB 2,000. Of the total 723 predictors included in the combined risk factor (n=5) and metabolites (n=718) model, most were retained in the sPTB model and least in the GDM model. At least four, of the five, established risk factors were retained in the risk factor models for all outcomes except SGA. A full list of the retained predictors is available in **Tables S5-S10**

**Table 2:** Number of predictors retained in each model. developed and tested in BiB 2,000 from total possible (n(%)). Percentages are rounded to the nearest whole number.

| Outcome | Model (retained predictors/total number of predictors possible [%]) |
| --- | --- |
| Gestational diabetes | Risk factor (4/5[80%]) |
|  | Metabolite (81/718 [11%]) |
|  | Combined (82/723 [11%]) |
| Gestational hypertension | Risk factor (4/5[80%]) |
|  | Metabolite (28/718 [4%]) |
|  | Combined (75/723 [10%]) |
| Pre-eclampsia | Risk factor (4/5 [80%]) |
|  | Metabolite (154/718 [21%]) |
|  | Combined (28/723 [4%]) |
| Small for gestational age | Risk factor (5/5 [100%]) |
|  | Metabolite (66/718 [9%]) |
|  | Combined (65/723 [8%]) |
| Large for gestational age | Risk factor (5/5 [100%]) |
|  | Metabolite (490/718 [68%]) |
|  | Combined (360/723 [50%]) |
| Spontaneous preterm birth | Risk factor (4/5 [80%]) |
|  | Metabolite (587/718 [83%]) |
|  | Combined (328/723 [45%]) |

We found little overlap between predictors retained in the combined risk factor and metabolite models across outcomes. There was an overlap of 41 predictors in the combined models for the GDM and LGA models, including 4-hydroxyglutamate, which was also retained for PE. There were five predictors retained in both the combined GDM and PE models, including 4’hydroxyglutamate and glycerol. There were 33 predictors retained for both LGA and SGA, including lanthionine, pipecolate, BMI, ethnicity, and parity.

In both BiB and POPs cohorts, discrimination for GDM, PE, LGA and SGA improved with the addition of metabolites to the risk factor only model (combined model). The combined model AUC’s were similar for both cohorts, with good discrimination for GDM and LGA (GDM - (AUC (95% CI)) BiB 0.76 (0.71,0.81) and POPs 0.76 (0.72,0.81); LGA - BiB 0.86 (0.80, 0.91) and POPs (0.76 (0.60, 0.92)). Discrimination was improved for the combined models compared to the risk factors for PE and SGA, but the AUC was modest (PE - BiB 0.68 (0.58, 0.78) and POPs 0.66 (0.60, 0.71); SGA – BiB 0.68 (0.63, 0.74)) and POPs (0.64 (0.59, 0.69)). In BiB, risk factors alone were the best predictor of GHT: 0.74 (0.68, 0.80) compared to 0.72 (0.66, 0.79) for the combined model. The GHT models could not be validated in POPs due to inadequate number of cases with metabolite data. Discrimination for sPTB was very poor for all models and in both cohorts, with a negligible improvement in the BiB 1,000 testing set with the addition of metabolites to the risk factors model (0.54 (0.40, 0.67) increasing to 0.56 (0.45, 0.67)) (**Figure 2**). In POPs, the AUC was 0.50 for both models (**Table S4**).

**Figure 2.**
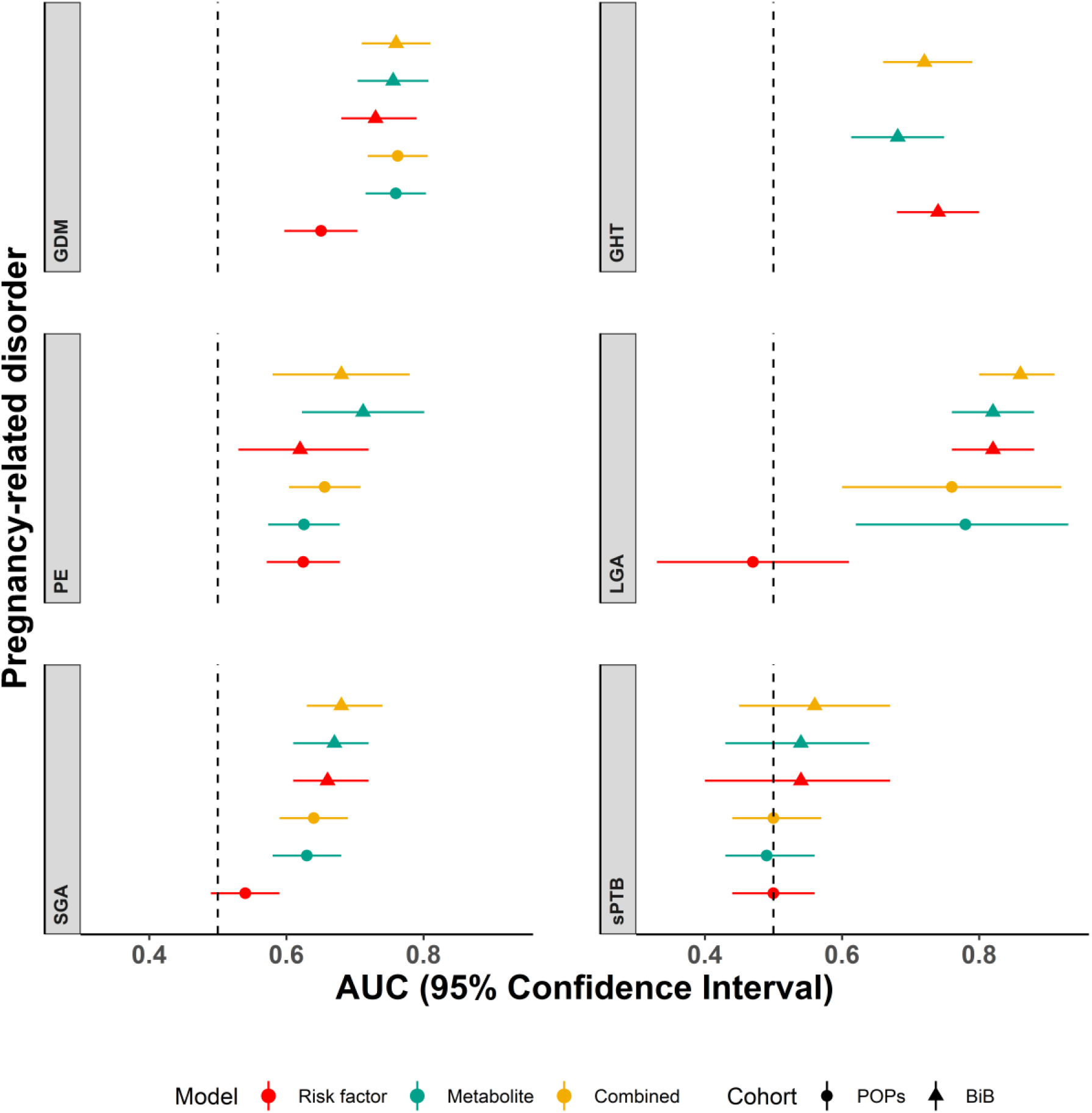
Predictive discrimination of models for each outcome. AUC and 95% confidence intervals are shown for established risk factor prediction models (red), metabolite models (green) and combined risk factor and metabolite models (yellow) trained in the Born in Bradford 2,000, tested in the Born in Bradford 1,000 (triangles) and external validation in the Pregnancy Outcome Prediction study (circles). POPs did not have sufficient data on gestational hypertension for validation. Abbreviations: BiB, Born in Bradford; POPs, Pregnancy Outcome Prediction study; GDM, gestational diabetes; GHT, gestational hypertension; PE, pre-eclampsia; SGA, small for gestational age; LGA, large for gestational age; sPTB, spontaneous preterm birth.

Calibration slopes showed good adherence of the predicted probabilities to the observed outcomes for the combined metabolite and risk factor models for GDM and SGA. Calibration was moderate for LGA. There was overestimation for GDM and underestimation for SGA and LGA as the intercepts show below (**Figure 3-5**). Calibration was attenuated for the models in POPs (**Figure S4-S6**). Calibration for the other outcomes was poor **(Figure S7-S8)**.

**Figure 3.**
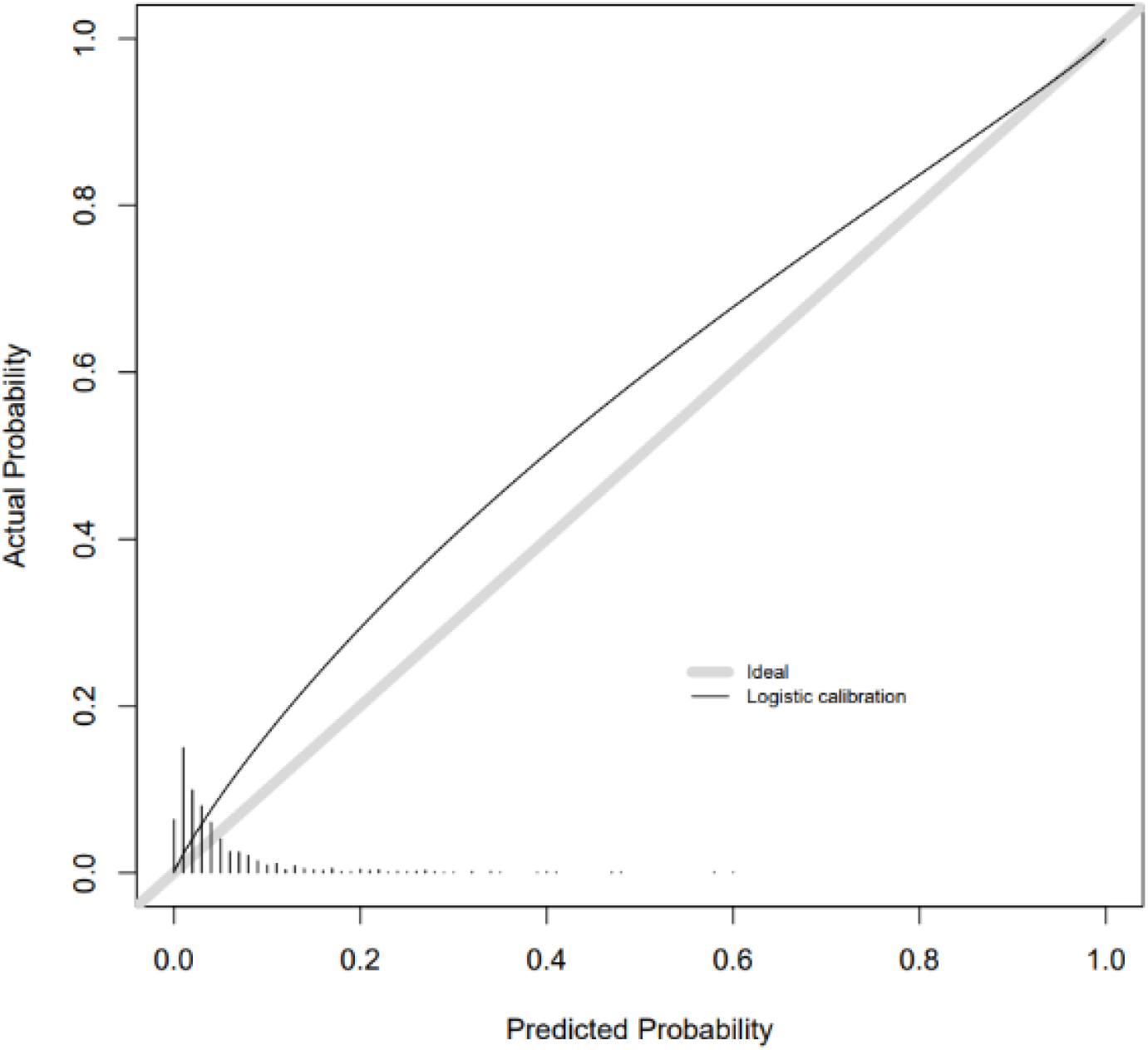
Calibration slope for GDM combined model in BiB 1,000 testing.

**Figure 4.**
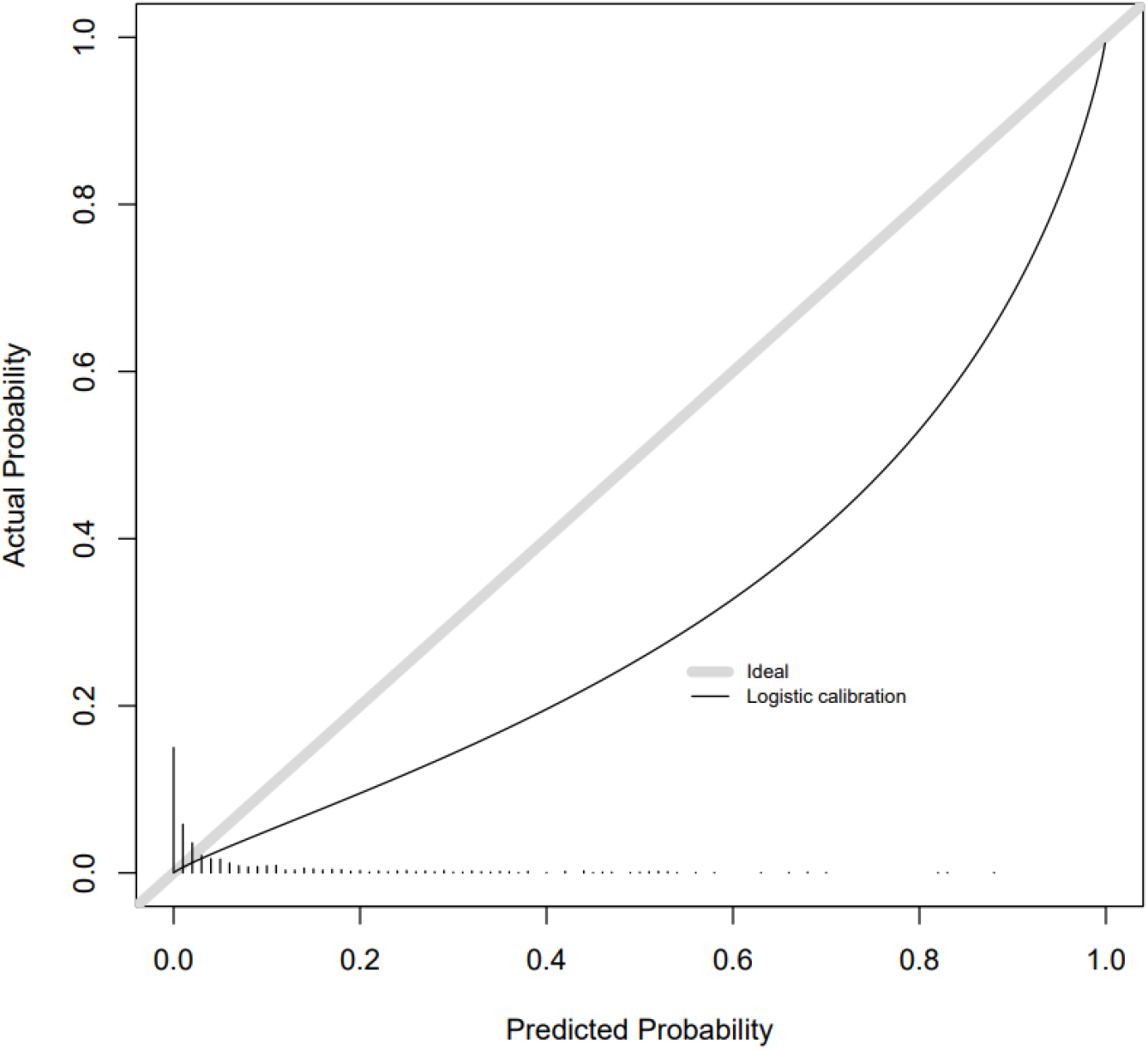
Calibration slope for LGA combined model in BiB 1,000 testing

**Figure 5.**
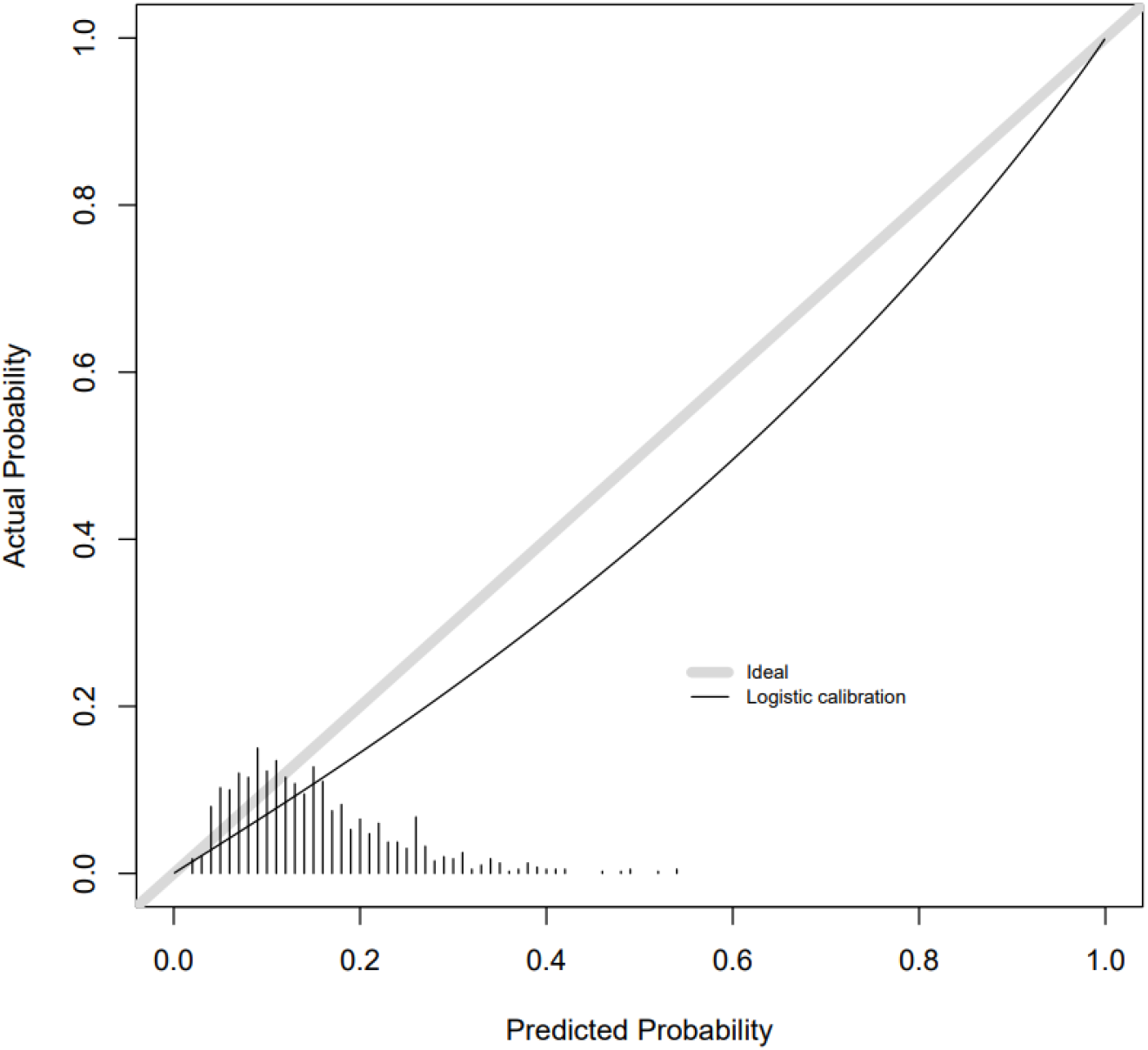
Calibration slope for SGA combined model in BiB 1,000 testing.

### Additional analyses

In an additional analysis of all PTB in BiB (141/1,441 cases in BiB 2,000 training and 38/915 cases in BiB 1,000 testing), results did not differ substantially from those for just sPTB (87/1,441 cases in BiB 2,000 training and 21/915 cases in BiB 1,000 testing), with poor discrimination (**Table S12**).

There were 828 women with ‘any’ pregnancy-related disorder in the BiB 2,000 (training) and 301 with ‘any’ pregnancy-related disorder in the BiB 1,000 (testing). The combined risk factors and metabolite model had very moderate discrimination, with an AUC for predicting ‘any’ pregnancy-related of 0.62 (0.58, 0.66). This is only a slight improvement on the discrimination for the clinical risk factor model 0.60 (0.56, 0.64). It did have good model calibration (**Figure S8**). Retained predictors include those retained in the models for GDM, PE and LGA including glutamate, 4-hydroxyglutamate, glycerol and the nucleotide 5,6-dihydrouracil (**Table S11**).

## Discussion

In this study we have shown improved prediction for GDM and LGA using a model that combines mass-spectrometry assessed metabolites and risk factor predictors, compared to established predictors only. We also found improved prediction of PE and SGA in the testing and external validation cohorts - but for these outcomes discrimination was modest. These models were tested and externally validated in two independent cohorts. The BiB cohort is largely socioeconomically deprived (68% living in an area of the highest quintile of socio-economic deprivation in England)(30), includes women of different ethnicities and is ∼40% nulliparous. POPs is more affluent (0% living in an area of the highest quintile of socioeconomic deprivation in England)(30), majority white women (∼95% in this study) and all women are nulliparous. Validation of the models trained and tested in BiB in POPs suggest the models are not influenced by overfitting, and are generalizable across diverse populations.

Calibration was good for the combined risk factor and metabolite model for GDM and SGA, and moderate for LGA. Calibration was consistently attenuated in the POPs validation. This would be expected, as the key determinant of calibration is the prevalence of the outcome in the underlying population and re-calibration (by modifying the model intercept based on population prevalence, whilst keeping all other parameters the same) is often used when comparing models between populations. We would therefore expect the calibration to be better in BiB 1,000, than POPs, as it is from the same underlying population as BiB 2,000. The marked differences between POPs and BiB in socioeconomic background, ethnicity, and parity would contribute to making outcome prevalence very different. The risk factor model validation of LGA and SGA performed particularly poorly in the POPs. This may be explained by participant characteristic differences between the two cohorts, such as ethnicity. Ethnicity is a good predictor of these outcomes in BiB. In observational analyses in POPs, there is no association between ethnicity and LGA and SGA, possibly due to very little variation in ethnicity in POPs (with 95% of women being White).

Due to known overlap in pregnancy-related disorders, we trained (BiB 2,000) and tested (BiB 1,000) prediction models for ‘any’ pregnancy-related disorder. The discrimination for the combined prediction model was modest, and not much improved from the risk factors model. This suggests that a single metabolomic prediction tool for ‘any’ pregnancy complication is unlikely to be feasible, despite the fact that calibration was good.

In our previous work, we found that NMR-derived metabolomics improves upon risk factors for prediction of GDM, LGA, SGA and combined PE and GHT (hypertensive disorders of pregnancy – HDP). We reported the best discrimination for GDM and LGA, and our findings here suggest that metabolites from different platforms are valuable for prediction of GDM and LGA(25). In previous work in POPs and BiB 1,000, we found that the amino acid 4-hydroxyglutamate was a novel predictor of PE and the metabolite ratio described above were a better predictor of fetal growth restriction/SGA than a biomarker ratio used in the diagnosis of PE (sFlt1:PlGF)(27, 30). The focus of these previous studies was to identify a small number of independently predictive metabolites to be used in a targeted assay. Moreover, BiB 2,000 was not included in the first of these two previous studies. The aim of this study was to evaluate predictive associations with a metabolomic profile using high dimensional statistical methods. The consistency in observations across the two distinct cohorts implies that some of these metabolites might be causal in determining the pregnancy complications studied. Consistent with our earlier published work, 4-hydroxyglutamate was one of the metabolites maintained in the combined model for PE in the new BiB 2,000 cohort. 4’hydroxyglutamate was also retained in the combined models for GDM, LGA and in the ‘any’ pregnancy-related disorder combined model. The lipid glycerol was a retained predictor for GDM, PE and the ‘any’ pregnancy-related disorder combined model. These represent potential markers for future study. Despite these individual metabolites being maintained in prediction models for more than one outcome, overall, there was relatively little overlap between the retained models where we had good prediction for different pregnancy-related disorders.

These findings suggest that metabolites (from different platforms) contribute to the improved prediction of GDM, LGA, SGA and PE, compared to models using only established maternal risk factors. Whilst some studies have also shown value in metabolomics for predicting PTB, most of those have small sample sizes, have not compared predictive ability to established risk factors or not externally validated findings(42-44). Our work, with external validation, suggests that PTB is not accurately predicted by either established risk factors, metabolomic profile, or the two combined.

### Strengths and limitations

Key strengths of this study are the internal and external (independent) validation and the exploration of a very large number of metabolite measures. We assessed discrimination and calibration of our prediction models and found using metabolomics improved performance compared to established risk factors for GDM, PE, LGA and SGA. MS has the advantage of giving greater coverage of the metabolome than other methods such as NMR, however it is more expensive (∼£150 a sample as opposed to ∼£25 a sample). Due to this, it is hard to find any other cohorts currently with MS data generated in pregnancy samples to strengthen our findings. None of these datasets represented random samples so future work should continue to validate these models in more general populations.

Despite trying to harmonise data between BiB and POPs there were some differences in outcomes between the two. POPs performed metabolomics on a subset of cases of PE with more severe PE, and excluded women with non-severe superimposed term PE from the case definition in the case-cohort design. There were only 12 cases of LGA in the POPs case-cohort as LGA was not one of the outcomes of interest in the original case-cohort analysis, and therefore the estimates have wide confidence intervals. In BiB 2,000 we included any PTB as cases, whereas in POPs the case definition included sPTB only. Therefore, we used sPTB in the main analysis which included both BiB and POPs. However, it is reassuring that results are consistent between the two studies. Whilst PE is the more severe form of HDP, GHT affects more women and is also associated with adverse perinatal and longer-term outcomes. Independent external validation of the GHT model is important but could not be performed in POPs since GHT was not included in the case-cohort design in this population.

Ideally, we would have a prediction tool that could be used as early as possible in pregnancy and be repeated throughout, for an updated risk assessment. This would allow women’s antenatal care to be tailored to their risk from early pregnancy. This was not possible in this study as we have only one sample taken at 26-28 weeks gestation in BiB. Furthermore, all three of the studies used here were selected, non-random samples (**Figure 1**), The models that we have found to improve prediction of pregnancy-related disorders need to be further tested on blood samples measured early in pregnancy and random samples of general populations of pregnant women.

## Conclusions

To conclude, our results suggest metabolomics combined with established risk factors can improve prediction of GDM and LGA. We validated our findings in an independent cohort. However, we acknowledge the need to validate these findings in a large independent sample of unselected pregnant women and examine their accuracy when measured earlier on in pregnancy. These findings show promise for the use of blood-derived metabolomics to improve prediction of common pregnancy complications. Further validation would support developing the tool using only the specific metabolites (rather than incurring the cost of the full panel) and testing its effectiveness in practice. Thus, our findings provide a promising evidence base for further research with the aim of being able to tailor antenatal care for women at risk of GDM and LGA.

## Data Availability

Data are available upon request from https://borninbradford.nhs.uk/research/how-to-access-data/. The POPs study data are available from G.C.S.S. upon reasonable request. Data requests will require a formal Data Transfer Agreement. Data are not publicly available due to the terms of the ethical approval.

## Abbreviations

GDM: gestational diabetes
PE: pre-eclampsia
GHT: gestational hypertension
LGA: large for gestational age
sPTB: spontaneous preterm birth
BMI: body mass index
BiB: Born in Bradford
POPs: Pregnancy Outcome Prediction study
AUC: Area under the curve
MS: mass spectrometry

## Acknowledgements

Born in Bradford is only possible because of the enthusiasm and commitment of the Children and Parents in BiB. We are grateful to all the participants, practitioners and researchers who have made Born in Bradford happen. The authors are extremely grateful to all the families who took part in this study, and the teams that make up BiB and POPs, which includes midwives, interviewers, computer and laboratory technicians, clerical workers, research scientists, volunteers, managers, receptionists, and nurses.

## Authors contributions

D.A.L and G.C.S.S. undertook funding acquisition and project administration. D.A.L., P.Y., C.R., and M.S. supervised N.M., where they conceived the study and developed the methodology. D.A.L, N.M., G.C.S.S., U.S., and K.T. performed the data curation. D.A.L., P.Y., C.R., M.S. and N.M. designed the experiment and analysis. N.M. and U.S. conducted the formal analysis and validation. D.A.L., G.C.S.S., P.Y., C.R., M.S., N.M., K.T. and U.S. drafted the original version of the manuscript. D.A.L., G.C.S.S., P.Y., C.R., M.S., N.M., K.T., U.S., V.F., B.H. and T.Y. provided data interpretation, critical review, editing and commentary to the revised versions of the manuscript. All authors have seen and approved the final versions of this manuscript.

